# Ongoing symptoms and functional impairment 12 weeks after testing positive to SARS-CoV-2 or influenza in Australia: an observational cohort study

**DOI:** 10.1101/2023.04.16.23288205

**Authors:** Matthew Brown, John Gerrard, Lynne McKinlay, John Marquess, Teneika Sparrow, Ross Andrews

## Abstract

**Objective:** In a highly vaccinated Australian population we aimed to compare ongoing symptoms and functional impairment 12 weeks after PCR-confirmed COVID-19 infection with PCR-confirmed influenza infection.

**Methods and Analysis:** The study commenced upon a positive PCR test for either COVID-19 or influenza in June 2022 during concurrent waves of both viruses. Participants were followed up 12 weeks later in September 2022 and self-reported ongoing symptoms and functional impairment. We conducted a multivariate logistic regression analysis, controlling for age, sex, First Nations status, vaccination status, and socio-economic profile.

**Results:** There were 2 195 and 951 participants in the COVID-19 and influenza-positive cohorts respectively. After controlling for potential predictor variables, we found no evidence to suggest adults with COVID-19 were more likely to have ongoing symptoms (21.4% vs 23.0%, aOR 1.18; 95% CI 0.92-1.50) or moderate to severe functional impairment (4.1% vs 4.4%, OR 0.81; 95% CI 0.55-1.20) at 12 weeks after their diagnosis than adults who had influenza.

**Conclusions:** In a highly vaccinated population exposed to the SARS-CoV-2 omicron variant, long COVID may manifest as a post-viral syndrome of no greater severity than seasonal influenza but differing in terms of the volume of people affected and the potential impact on health systems. This study underscores the importance of long COVID research featuring an appropriate comparator group.

**What is already known on this subject?:** Post-acute infection syndromes are associated with a range of illnesses, including COVID-19 and influenza. “Long COVID” may pose a risk to health systems.

**What are the new findings?:** In a highly vaccinated population whose primary exposure has been to the Omicron variant, the rates of ongoing symptoms and moderate to severe functional impairment at 12 weeks after COVID-19 are no different to influenza.

**How might these results change the focus of research or clinical practice?:** The public health impact arising from long COVID may not stem from severity, but from volume. We do not dismiss the validity of long COVID but recommend an appropriate comparator group when researching this condition.

## INTRODUCTION

Long COVID, now variously described as post-COVID condition, post-COVID syndrome, and post-acute sequelae of COVID-19,[1] has been described as the pandemic’s next challenge to Governments and global health systems.[2]

The World Health Organization (WHO) definition of long COVID includes ongoing symptoms three months after infection.[1] Others use different definitions and time periods, or offer refinements to the WHO definition.[3] A diagnosis requires the elimination of other possible explanations and must consider over 200 potential and highly heterogenous symptoms.[4-7] Many of these occur commonly in the general population,[8] and persistent sequelae have been associated with past viral outbreaks and pandemics.[9]

Research has noted the available evidence is frequently low quality, prone to bias and often lacking an appropriate comparator group.[10] While many studies focused on SARS-COV-2 variants in unvaccinated populations early in the pandemic, relatively few have examined long COVID in vaccinated populations in the omicron era.[11] As a result, the impact of long COVID in Australia’s context remains largely unquantified.

In Australia, the first wave of the omicron variant commenced in late 2021 when over 90% of the population was double vaccinated against COVID-19.[12] Until that time, the Australian state of Queensland was relatively COVID-19 naïve. This study sought to understand the potential impact of long COVID on the Queensland population and to inform the heath system response. Our primary aims were to determine whether a cohort of adults aged 18 years or more who tested RT-PCR positive to either COVID-19 or influenza were more likely to have ongoing symptoms or moderate to severe functional limitations 12 weeks later if they had COVID-19 (i.e. long COVID) in comparison to influenza.

## MATERIALS AND METHODS

### Study design and participants

We conducted an observational cohort study among individuals aged 18 years and older who tested RT-PCR positive for COVID-19 or influenza between 12 and 25 June 2022 in Queensland. The primary exposure of interest was a diagnosis of COVID-19 versus influenza. Laboratory reporting for these two conditions is mandated upon test request under Queensland’s public health legislation,[13] with this data recorded in the Queensland Department of Health’s Notifiable Conditions System (NoCS). The date range corresponded with the commencement of a COVID-19 wave and the seasonal influenza A peak in Queensland.[14] An additional group consisting of individuals who tested RT-PCR negative to both conditions but met all other selection criteria was recruited as a secondary outcome comparator (data not shown here).

The primary outcomes of interest were a) ongoing symptoms and b) moderate to severe functional limitation. These were assessed through a self-administered questionnaire between 12 and 22 September 2022 (12 weeks post each participant’s PCR test result).

Our report conforms with the STrengthening the Reporting of OBservational studies in Epidemiology (STROBE) guidelines.[15]

### Procedures

Each eligible participant’s mobile telephone number, first name and date of their PCR test was extracted from NoCS. The data were cleaned, collated and each record was assigned a unique identifier to enable matching with subsequent extraction of relevant predictor variables from NoCS. Participants were sent a text message and the questionnaire using the Whispir communications platform on weeknights between 12 and 22 September 2022. One short reminder text message was sent the following day. The survey closed 48 hours after the initial text message was sent.

The introductory text was addressed to the individual’s first name and indicated the message was from Queensland Health to follow-up a PCR test for [COVID-19 or influenza] on the relevant date in June 2022.

Respondents were asked if they had ongoing symptoms that related to their initial PCR test, and if so, the degree of functional impairment. The questionnaire is provided as supporting information. The Post-COVID Functional Status (PCFS) tool was used as the basis for grading,[16] because of its demonstrated utility to discriminate between a range of symptom and functional domains common in post-viral syndromes.[17] Modifications were made to provide the brevity and clarity required in a mobile phone-based questionnaire, and to include specific reference influenza where applicable rather than solely to COVID-19. The grading scores were: no symptoms or no limitations to daily activities (Grade 0); ongoing symptoms but no effect on daily activities (Grade 1); ongoing symptoms and occasional limitations on daily activities (Grade 2); ongoing symptoms that limit all daily activities, with an ability to take care of oneself without assistance (Grade 3); and ongoing symptoms resulting in severe limitations in everyday life, and with a dependency on another person for care (Grade 4).[16] Consistent with other studies, participants with a PCFS score of 3 or 4 were defined as having moderate to severe functional limitations.[17]

Potential predictor variables were ascertained from demographic information routinely recorded in NoCS, COVID-19 vaccination records, and the Index of Relative Socio-economic Advantage and Disadvantage (IRSAD) produced from the Australian Bureau of Statistics.[18] The IRSAD score is ranked in deciles from 1-10 based on economic, educational and social domains, with 1 being the most disadvantaged and 10 being the most advantaged.

Eligible participants were all those persons recorded on the NoCS system with either condition confirmed from a specimen collected within the specified date range, and a valid Australian mobile telephone number. Only one individual was eligible per mobile number. Participants were excluded from further analyses if they self-reported at the 12-week follow-up that they had symptoms or a new illness that was unrelated to the reasons for their initial PCR test.

### Statistical analysis

The primary outcomes (ongoing symptoms, moderate to severe functional impairment) and predictor variables were expressed as proportions. We used multivariable logistic regression analysis for each primary outcome as the dependant variable, with the influenza PCR positive individuals as the referent group. Predictor variables assessed for inclusion were age, sex, First Nations status, COVID-19 vaccine dose (<3 doses (referent) versus 3 or more) at the time of testing, vaccine recency at the time of testing (<6 months (referent) versus 6 or more months since last COVID-19 vaccine dose), and the IRSAD score for the participant’s residential address at statistical area 1 with the most disadvantaged quintile as the referent group. Each of the potential predictor variables were assessed against the relevant primary outcome of interest in univariable analysis using chi-square tests. Those that were significant at the 10% level were included in the multivariable logistic regression model as potential predictors and removed in stepwise fashion if not significant at the 5% level. Adjusted odds ratios (*a*ORs) and 95%CI were reported.

### Ethics approval and study registration

The study was approved by the Metro South Health Research Ethics Committee (HREC/2022/QMS/88587) and the Queensland Office of Precision Medicine and Research (SSA/2022/QHC/88587). Participants were given information on the study prior to commencing the questionnaire and provided electronic consent by electing to participate. The study was registered with the Australian New Zealand Clinical Trials Registry (ACTRN12623000041651).

### Patient and public involvement

This study was undertaken during a declared public health emergency to urgently ascertain the potential health burden facing a state health system. It was undertaken by the Queensland Government Department of Health under section 83 of the Queensland Public Health Act 2005. Participants were not involved in the design, conduct, or reporting of this rapid response research. We aim to engage the public through dissemination of the findings through the Queensland Department of Health.

## RESULTS

There were 3 146 participants in the study cohort at a ratio of 2.3:1 for individuals diagnosed with COVID-19 compared to influenza (Figure 1). Overall, there were 19 758 individuals who had tested PCR positive to either condition during the 2-week eligibility period and 12 932 individuals potentially contactable through a unique valid mobile telephone number. Within each cohort (COVID-19 vs influenza), the eligible proportion (65% vs 67%), the consent rate (27% vs 26%) and post enrolment exclusions (8% vs 9%) were similar. Compared with non-participants for each cohort, there was a relatively higher proportion of females, persons aged ≥?50 years, 3 vaccine dose recipients, recent vaccinees and less disadvantaged individuals among the study participants (Table 1).

**Table 1.** Baseline characteristics of participants and non-participants within the eligible cohort groups. ^1^ IRSAD: Index of Relative Socio-economic Advantage and Disadvantage where IRSAD 1 = most disadvantaged and IRSAD 10 = most advantaged.

|  | Influenza PCR Positive |  |  |  |  | Covid-19 PCR Positive |  |  |  |  |
| --- | --- | --- | --- | --- | --- | --- | --- | --- | --- | --- |
| Characteristic | Non-participant |  | Participants |  |  | Non-participant |  | Participants |  |  |
|  | (n=3127) |  | (n=951) |  |  | (n=6660) |  | (n=2195) |  |  |
|  | n | % | n | % | Diff. | n | % | n | % | Diff. |
| Female | 1857 | 59.4% | 676 | 71.1% | 11.7% | 3570 | 53.6% | 1351 | 61.6% | 8.0% |
| Age at diagnosis |  |  |  |  |  |  |  |  |  |  |
| 18-39 years | 1794 | 57.4% | 426 | 44.8% | -12.6% | 2674 | 40.2% | 570 | 26.0% | -14.2% |
| 40-49 years | 520 | 16.6% | 171 | 18.0% | 1.4% | 1184 | 17.8% | 383 | 17.5% | -0.3% |
| ≥50 years | 810 | 25.9% | 354 | 37.2% | 11.3% | 2802 | 42.1% | 1241 | 56.6% | 14.5% |
| First Nations People | 195 | 6.2% | 35 | 3.7% | -2.5% | 228 | 3.8% | 38 | 1.8% | -2.0% |
| Covid-19 vaccination |  |  |  |  |  |  |  |  |  |  |
| ≥3 doses | 1384 | 44.3% | 604 | 63.5% | 19.2% | 2689 | 67.1% | 1220 | 83.2% | 16.1% |
| ≥6 months since last dose | 1038 | 39.7% | 286 | 33.4% | -6.3% | 1273 | 31.8% | 338 | 23.1% | -8.7% |
| Socio-economic status<sup>1</sup> |  |  |  |  |  |  |  |  |  |  |
| IRSAD score=1 | 225 | 10.6% | 58 | 8.5% | -2.1% | 478 | 10.0% | 102 | 6.2% | -3.8% |
| IRSAD score=2 | 222 | 10.5% | 39 | 5.7% | -4.8% | 505 | 10.5% | 138 | 8.3% | -2.2% |
| IRSAD score=3 | 184 | 8.7% | 44 | 6.5% | -2.2% | 486 | 10.1% | 174 | 10.5% | 0.4% |
| IRSAD score=4 | 192 | 9.1% | 68 | 10.0% | 0.9% | 373 | 7.8% | 136 | 8.2% | 0.4% |
| IRSAD score=5 | 208 | 9.8% | 62 | 9.1% | -0.7% | 457 | 9.5% | 142 | 8.6% | -0.9% |
| IRSAD score=6 | 193 | 9.1% | 62 | 9.1% | 0.0% | 467 | 9.7% | 181 | 10.9% | 1.2% |
| IRSAD score=7 | 208 | 9.8% | 66 | 9.7% | -0.1% | 431 | 9.0% | 162 | 9.8% | 0.8% |
| IRSAD score=8 | 223 | 10.5% | 77 | 11.3% | 0.8% | 540 | 11.3% | 157 | 9.5% | -1.8% |
| IRSAD score=9 | 211 | 10.0% | 87 | 12.8% | 2.8% | 525 | 10.9% | 231 | 14.0% | 3.1% |
| IRSAD score=10 | 250 | 11.8% | 119 | 17.4% | 5.6% | 536 | 11.2% | 232 | 14.0% | 2.8% |

**Figure 1.**
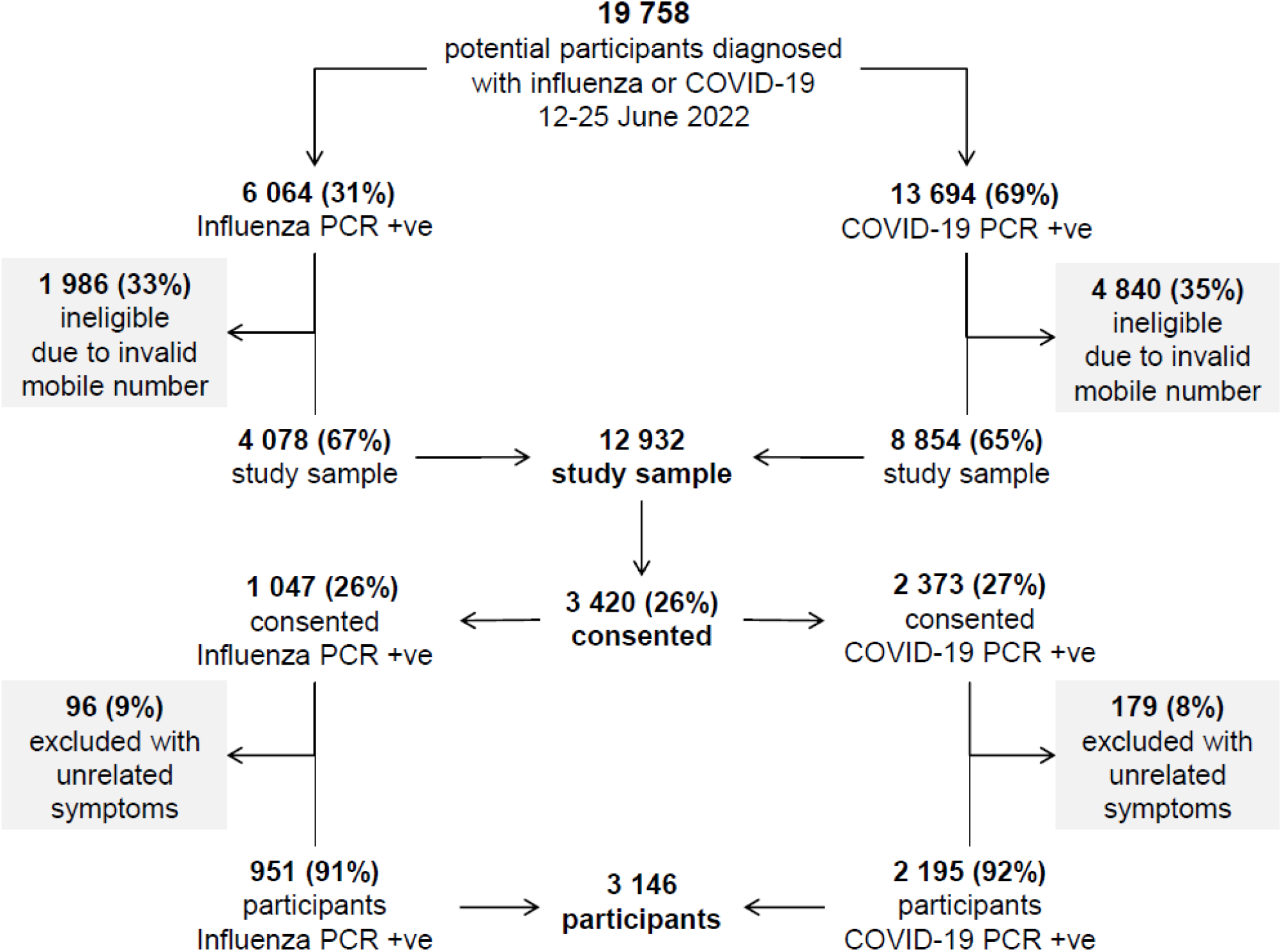
Flow diagram of study sample and participant groups.

Ongoing symptoms were reported by 21.4% of participants who had been COVID-19 positive when tested 12 weeks earlier and by 23.0% of those who had been influenza positive. After adjusting for all the included covariables (Table 2) the aOR was 1.18 (95%CI 0.92 to 1.50). Moderate to severe functional impairment was identified among 4.1% of participants who had been COVID-19 positive when tested 12 weeks earlier and by 4.4% of those who had been influenza positive. After adjusting for all the included covariables (Table 2) the aOR was 0.81 (95%CI 0.55 to 1.12). Figure 2 shows covariables, referents and aORs for both ongoing symptoms and moderate to severe functional impairment. Predictors of self-reported ongoing symptoms at 12 weeks were being female rather than male and being the most disadvantaged on the IRSAD scale when compared with the midpoint on the IRSAD scale. Predictors of severe to moderate functional impairment were being a First Nations person and being of older age.

**Table 2.** Proportion of participants with ongoing symptoms and moderate to severe functional impairment.

| Primary outcome | Covid-19 PCR Positive | Influenza PCR Positive | Odds Ratio | P>z | [95% Conf. Interval] |  |
| --- | --- | --- | --- | --- | --- | --- |
| Ongoing symptoms | 21.4% (469/2195) | 23.0% (214/951) | 1.18 | 0.19 | 0.92 | 1.50 |
| Predictor variables |  | Female | 1.65 | 0.00 | 1.29 | 2.13 |
|  |  | IRSAD score=1-2 | referent | referent | referent | referent |
|  |  | IRSAD score=3-4 | 0.76 | 0.17 | 0.52 | 1.13 |
|  |  | IRSAD score=5-6 | 0.66 | 0.03 | 0.45 | 0.97 |
|  |  | IRSAD score=7-8 | 0.68 | 0.05 | 0.47 | 1.00 |
|  |  | IRSAD score=9-10 | 0.82 | 0.25 | 0.58 | 1.16 |
|  |  | ≥ 3 COVID 19 vaccine doses | 0.73 | 0.02 | 0.56 | 0.96 |
|  |  | Constant | 0.29 | 0.00 | 0.19 | 0.43 |
| Moderate to severe functional impairment | 4.1% (90/2195) | 4.4% (42/951) | 0.81 | 0.29 | 0.55 | 1.20 |
| Predictor variables |  | Age 18 to 39 years | referent | referent | referent | referent |
|  |  | 40 to 49 years | 2.01 | 0.02 | 1.13 | 3.58 |
|  |  | ≥50 years | 2.17 | 0.00 | 1.35 | 3.50 |
|  |  | First Nations | 3.28 | 0.00 | 1.52 | 7.06 |
|  |  | Constant | 0.03 | 0.00 | 0.02 | 0.04 |

**Figure 2.**
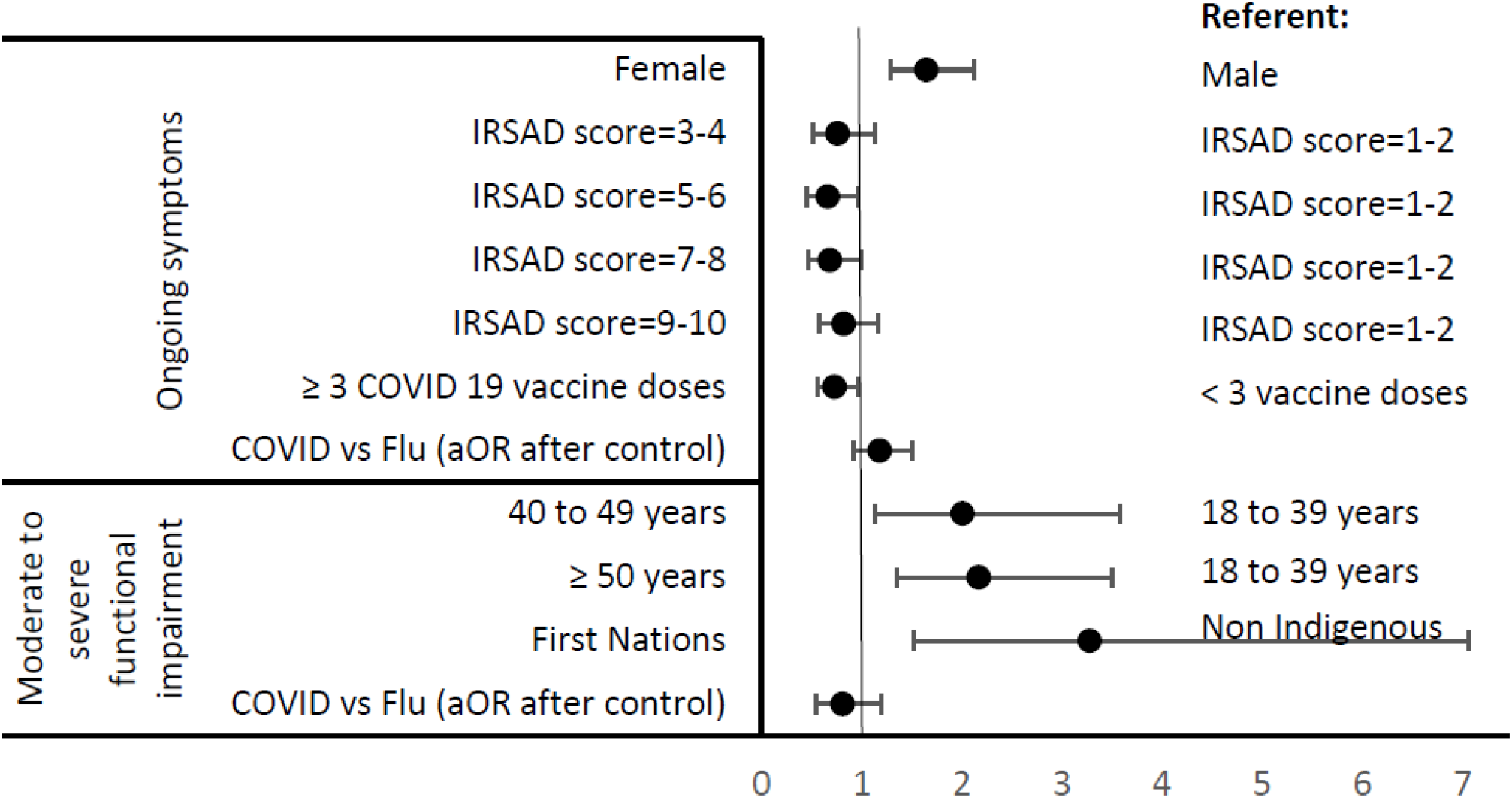
ongoing symptoms and moderate to severe functional impairment – adjusted odds ratios for COVID-19 compared with influenza.

## DISCUSSION

We found that adults who tested PCR positive to the SARS-CoV-2 omicron variant were no more likely to have ongoing symptoms at 12 weeks after their test (21.4%) than adults who tested PCR positive to influenza (23.0%. aOR 1.18; 95% CI 0.92–1.50). Similarly, we found no evidence to suggest that adults diagnosed with COVID-19 were more likely to have moderate to severe functional impairment at 12 weeks after their test (4.1%) than adults who tested PCR positive to influenza (4.4%. OR 0.81; 95% CI 0.55–1.20). Being female and socio-economically disadvantaged were predictors of ongoing symptoms while being a First Nations person or an older person were predictors of moderate to severe functional limitations.

To our knowledge, this is the first observational cohort study in a highly vaccinated population that directly compares ongoing symptoms and degree of functional impairment after infection by the SARS-CoV-2 omicron variant with influenza. Our study was undertaken during the 2022 peak of seasonal influenza in Queensland and at the commencement of a third wave of the omicron variant when approximately 40% of adults had been vaccinated for influenza and over 93% of adults had received at least two vaccinations for COVID-19.[12,19]

Our findings suggest that, in a highly vaccinated population, the odds of having long COVID arising from the omicron variant are no greater than the odds of having a post-viral illness following seasonal influenza. The finding does not discredit long COVID as a health issue given the significant volume of COVID-19 infections when compared with seasonal influenza, noting that there was a 38-fold difference in reported case numbers (1,606,171 COVID-19 vs 42,338 influenza) between 1 January and 9 September 2022 in Queensland.[14,20] The substantial difference in the incidence of infection with a pandemic virus like SARS-CoV-2 and an endemic virus like seasonal influenza may make it appear that post-viral syndromes are unusually common with the novel pathogen, as cases in the community may be high without individual risk being greater.

We acknowledge higher proportions of females, older persons, vaccinees, and more advantaged individuals among the study participants, and that those who were fully recovered may have been less likely to respond.[21] Others have shown use of a mobile phone survey when compared with face-to-face data collection increases the adjusted odds ratio for self-reporting long COVID by 30%.[22]

Our study was unable to consider participant’s pre-existing health conditions, nor consider the severity of COVID or influenza infection. Participants who were hospitalised or in intensive care were not identifiable within the cohort. Emerging Australian research shows a low prevalence of long COVID in vaccinated adults following Omicron infection, albeit with a difference between adults who were hospitalised (1.9%) and not hospitalised (0.09%).[23] We do not discern if COVID-19 and influenza differ in specific symptom domains, including in areas of functional impact. However, we do discriminate between different levels of fatigue and functional performance, using consistent methods to those used where the PCFS grade 3–4 scale has shown poorer functional outcome, more fatigue and poorer quality of life.[17] We consider the PCFS grades 3–4 scale allows identification of the population of interest when considering the potential for impact on Queensland’s health system.

Previous reports have noted the high rate of typical long COVID symptoms amongst those recovering from influenza.[24] The high number and heterogenous nature of potential long COVID symptoms also underscores the importance of future studies featuring a concurrent control group. Our study supports recent literature suggesting that, in a highly vaccinated population, the SARS-CoV-2 omicron variant does not result in a significant burden of long COVID.[25] In this context, long COVID manifests at the population level as a post-viral syndrome of no greater severity than seasonal influenza but differing in terms of the volume of people affected and the potential impact on health systems.

## Data Availability

Requests for deidentified data associated with this research should be sent to the corresponding author after publication of the paper in the form of a formal data request which outlines the proposed use of this data and ensures appropriate attribution to this research.

## Data sharing

Because of data confidentiality provisions under Queensland public health legislation, individual data from Queensland Health’s Notifiable Conditions System will not be shared publicly. We can share the research protocol and survey questions. Requests for deidentified data associated with this research should be sent to the corresponding author after publication of the paper in the form of a formal data request which outlines the proposed use of this data and ensures appropriate attribution to this research.

## Author contributions

LM, JG, and MB conceptualised the study. MB designed the methodology and wrote the protocol and draft manuscript, with input from JG and LM. RA oversaw the data curation and extraction from the Notifiable Conditions System, led the formal analysis, and described this in the paper’s methodology and results. JM established the data curation protocols for records from the Notifiable Conditions System. TS provided the technical design, development, implementation and collation of the online survey, data cleansing and collation before and after the survey distribution. All authors critically revised and edited the manuscript, confirm they had full access to all the data in the study, and accept responsibility to submit for publication.

## Statements

The Corresponding Author has the right to grant on behalf of all authors and does grant on behalf of all authors, an exclusive licence (or non exclusive for government employees) on a worldwide basis to the BMJ Publishing Group Ltd to permit this article (if accepted) to be published in BMJ editions and any other BMJPGL products and sublicences such use and exploit all subsidiary rights, as set out in our licence.

All authors have completed the Unified Competing Interest form (available on request from the corresponding author) and declare: no support from any organisation for the submitted work; no financial relationships with any organisations that might have an interest in the submitted work in the previous three years, no other relationships or activities that could appear to have influenced the submitted work.

I, as lead author (the manuscript’s guarantor), affirm that the manuscript is an honest, accurate, and transparent account of the study being reported; that no important aspects of the study have been omitted; and that any discrepancies from the study as planned have been explained.

