## supporting information for "Ongoing symptoms and functional impairment 12 weeks after testing positive to SARS-CoV-2 or influenza in Australia: an observational cohort study"

Queensland Health Questionnaire

| | Content displayed | Selection option | Subsequent direction / advice | Alignment to PCFS Grade |
|---|---|---|---|---|
| Q1 | Do you understand the participant information and wish to proceed? | ☐ Yes | To Frame and Q2 | |
| | | ☐ No | Unfortunately we are unable to continue. If you have any questions, please call 13HEALTH (13 43 25 84) [End] | |
| [Frame] When answering the questions, please think about how your life and health are now, compared with before you got the symptoms that resulted in the [COVID-19* / influenza*] test on @@Date@@. *[*appropriate test displayed]* | | | | |
| Q2 | Has your health returned to the level it was before you needed to do the test on @@Date@@? | ☐ Yes - because I no longer have the symptoms that resulted in my test. | To Q5 | PCFS Grade = 0 |
| | | ☐ No - because I am still experiencing ongoing symptoms. | To Q3 | See Q3 |
| | | ☐ No - because I am currently unwell with unrelated symptoms or new illness. | If you are concerned about these new symptoms, please see your doctor or call 13HEALTH (13 43 25 84). (To Q5) | N/A |
| Q3 | Have your levels of everyday activity returned to where they were before your test on @@Date@@? | ☐ Yes | Q5 | PCFS Grade = 1 |
| | For example, are you capable of working the same work hours as before the test; can you look after yourself to the same level as before the test; can you undertake the same usual activities (like | ☐ No | Q4 | See Q4 |

| | | | | |
|---|---|---|---|---|
| | exercise/sport, duties at home) as before the test? | | | |
| Q4 | Please select one option below that best describes your current levels of health and everyday activity. | ☐ Because of my ongoing symptoms, I occasionally need to reduce, spread out or avoid some usual daily activities. However, I can perform these activities without any assistance. | Q5 | PCFS = 2 |
| | | ☐ Because of my ongoing symptoms, I can no longer perform all my usual activities. However, I can take care of myself without any assistance. | | PCFS = 3 |
| | | ☐ Because of my ongoing symptoms, I have severe restrictions on my usual activities. I now cannot take care of myself and am dependent upon another person (eg nurse, family member). | | PCFS = 4 |
| Q5 | Can Queensland Health contact you again to understand more about your recovery? | ☐ Yes | Submit Button [End] | |
